# Association of machine learning-derived measures of body fat distribution with cardiometabolic diseases in >40,000 individuals

**DOI:** 10.1101/2021.05.07.21256854

**Authors:** Saaket Agrawal, Marcus D. R. Klarqvist, Nathaniel Diamant, Takara L. Stanley, Patrick T. Ellinor, Nehal N. Mehta, Anthony Philippakis, Kenney Ng, Melina Claussnitzer, Steven K. Grinspoon, Puneet Batra, Amit V. Khera

## Abstract

**Background:** The clinical implications of BMI-independent variation in fat distribution are not fully understood.

**Methods:** We studied MRI imaging data of 40,032 UK Biobank participants. Using previously quantified visceral (VAT), abdominal subcutaneous (ASAT), and gluteofemoral (GFAT) adipose tissue volume in up to 9,041 to train convolutional neural networks (CNNs), we quantified these depots in the remainder of the participants. We derived new metrics for each adipose depot – fully independent of BMI – by quantifying deviation from values predicted by BMI (e.g. VAT adjusted for BMI, VATadjBMI) and determined associations with cardiometabolic diseases.

**Results:** CNNs based on two-dimensional projection images enabled near-perfect estimation of VAT, ASAT, and GFAT, with r^2^ in a holdout testing dataset (r^2^ = 0.978-0.991). Using the newly derived measures of local adiposity – residualized based on BMI – we note marked heterogeneity in associations with cardiometabolic diseases. Taking presence of type 2 diabetes as an example, VATadjBMI was associated with significantly increased risk (odds ratio per standard deviation increase (OR/SD) 1.49; 95%CI: 1.43-1.55), while ASATadjBMI was largely neutral (OR/SD 1.08; 95%CI: 1.03-1.14) and GFATadjBMI conferred protection (OR/SD 0.75; 95%CI: 0.71-0.79). Similar patterns were observed for coronary artery disease.

**Conclusions:** Deep learning models trained on a simplified MRI input enable near perfect quantification of VAT, ASAT, and GFAT. For any given BMI, measures of local adiposity have variable and divergent associations with cardiometabolic diseases.

## INTRODUCTION

Obesity is a leading threat to global public health, with afflicted individuals at increased risk of cardiovascular events, type 2 diabetes, cancer, and severe COVID-19 infection.^1–3^ Recent projections suggest that obesity – defined by body mass index (BMI) of at least 30 kg/m^2^ – will affect more than half of the U.S. adult population as soon as 2030.^4,5^

Although individuals with increased BMI tend to have higher risk of adverse outcomes on average, previous studies have suggested considerable heterogeneity.^6–9^ These studies have sought to define markers of “metabolic health” – such as measures of insulin resistance or waist circumference – as drivers of “within BMI-group variation” in cardiometabolic risk.^9–11^

Variation in fat distribution is a potential unifying explanation for cardiometabolic risk differences between two individuals with the same BMI.^12,13^ Prior studies have suggested that various fat depots have differing metabolic programs, with visceral adipose tissue (VAT) most strongly associated with cardiometabolic risk – but have potential limitations.^14–16^ First, most imaging studies to date have been cross-sectional and relatively small – especially those utilizing the gold-standard MRI modality – limiting ability to assess for depot-specific effects across age, sex, and BMI subgroups.^12,17–21^ Deep learning models trained on a small set of labeled images and subsequently applied to a larger set of unlabeled images may be one strategy to increase sample size if models were sufficiently predictive. Second, gluteofemoral adipose tissue (GFAT) – which may serve as an adaptive energy storage depot and a possible modifier of insulin resistance – has not been quantified in most previous imaging studies.^18–22^ Third, fat depot volumes tend to be highly correlated with both BMI and one another, making it challenging to isolate depot-specific associations with disease.^23^

In this study, we downloaded raw MRI imaging data from 40,032 participants of the UK Biobank and tested the hypothesis that deep learning models can be used to precisely quantify three fat depot volumes: VAT, abdominal subcutaneous adipose tissue (ASAT), and GFAT. We derived new measures for local adiposity burden, each fully independent of BMI, and note significant heterogeneity in risk conferred: VAT adjusted for BMI (VATadjBMI) associated with increased risk of type 2 diabetes and coronary artery disease, ASATadjBMI largely risk-neutral, and GFATadjBMI conferring protection.

## METHODS

### Study population

The UK Biobank is an observational study that enrolled over 500,000 individuals between the ages of 40 and 69 years between 2006 and 2010, of whom 43,521 underwent body MRI imaging between 2014 and 2020 as part of an imaging substudy.^24,25^ Images were acquired using the Dixon method, an MRI sequence that can be used to isolate fat signal from water signal.^26^ Each participant’s MRI data consisted of 244 axial slices acquired from the neck to the knees in four sequences: in-phase, out-of-phase, fat-only, and water-only. After exclusion of 3,489 (8.0%) of imaging scans based on technical problems or artifacts (**Supplementary Methods**), 40,032 participants remained for analysis. This analysis of data from the UK Biobank was approved by the Mass General Brigham institutional review board and was performed under UK Biobank application #7089.

### Machine learning to measure fat depot volumes

Among the 40,032 individuals with MRI imaging data available, a subset had visceral adipose tissue (VAT) volume, abdominal subcutaneous adipose tissue (ASAT) volume, and total adipose tissue (TAT) volume between the top of vertebrae T9 and the bottom of the thigh muscles, quantified and made available as previously described (N=9,040, 9,041, 7,754 participants, respectively).^20,21,27,28^ Gluteofemoral adipose tissue (GFAT) volume was derived by computing the difference between TAT and the sum of VAT and ASAT (**Supplementary Methods**). For each participant, we transformed three-dimensional MRI images into two-dimensional coronal and sagittal projections by computing the mean intensity projection in each orientation. For example, a given pixel on a coronal two-dimensional projection represents the mean intensity across all pixels making up a line oriented in the anterior-posterior direction perpendicular to the coronal plane. This procedure was done for the fat-only and water-only MRI sequences and the resulting images were jointly used as the imaging input for a given participant.

Individuals with previously quantified fat depot volumes were randomly split into 80% for training and a 20% holdout sample for testing. Five-fold cross-validation within the 80% training data was used to estimate error. Convolutional neural networks using two-dimensional coronal and sagittal MRI projections as inputs were evaluated in the 20% holdout sample, and subsequently used to predict fat depot volumes in the remaining participants with raw MRI imaging data but without labels. More comprehensive descriptions of the deep learning modeling and quality control – and the open-source code repository for data ingestion – are provided in the **Supplementary Methods**.

### Cardiometabolic disease definitions

Type 2 diabetes was defined on the basis of ICD-10 codes, self-report during a verbal interview with a trained nurse, use of diabetes medication, or a glycated hemoglobin greater than or equal to 6.5% before the date of imaging. Coronary artery disease was defined as myocardial infarction, angina, coronary revascularization, or death from coronary causes as determined on the basis of ICD-10 codes, ICD-9 codes, OPCS-4 surgical codes, nurse interview, and national death registries.

### Statistical analysis

We generated BMI-adjusted fat depot measurements by computing residuals from sex-specific linear regression models using BMI as the predictor and fat depot volume as the outcome, analogous to prior studies of waist-hip ratio adjusted for BMI.^29,30^ Logistic regression models were used to test the association of BMI-adjusted fat depot measurements with prevalent disease. To compute odds ratios across age, sex, and BMI subgroups, models were adjusted for age, sex (except in sex subgroup analyses), BMI, the other two fat depots (e.g. ASATadjBMI and GFATadjBMI for VATadjBMI), and MRI imaging center. Cox proportional-hazard models with the same covariates were used to test associations of BMI-adjusted fat depots with incident type 2 diabetes and coronary artery disease events. Finally, we used sex-stratified logistic regression to determine the gradient in probability of prevalent disease across clinical BMI categories. These models included interaction terms with body mass index along with the previously noted covariates and were standardized to the median of all predictor variables (except for MRI imaging center variable, where the mean was used) within each population.^4^ Effect sizes are reported per sex-specific standard deviation.

All analyses were performed with the use of R software, version 3.6.0 (R Project for Statistical Computing).

## RESULTS

Among 40,032 participants with MRI data available, the mean age was 65 years, 51% were women, and 97% were white (**Table 1**). Mean BMI was 26.1 kg/m^2^ among women and 27.1 kg/m^2^ among men, and mean waist-hip ratio (WHR) was 0.82 among women, and 0.94 among men. 1,901 individuals had been diagnosed with type 2 diabetes (4.7%) and 1,956 with coronary artery disease (4.9%) at the time of imaging assessment. VAT, ASAT, and GFAT volumes were previously quantified in 9,040, 9,041, and 7,754 participants, respectively.^20,21,27,28^

**TABLE 1.**
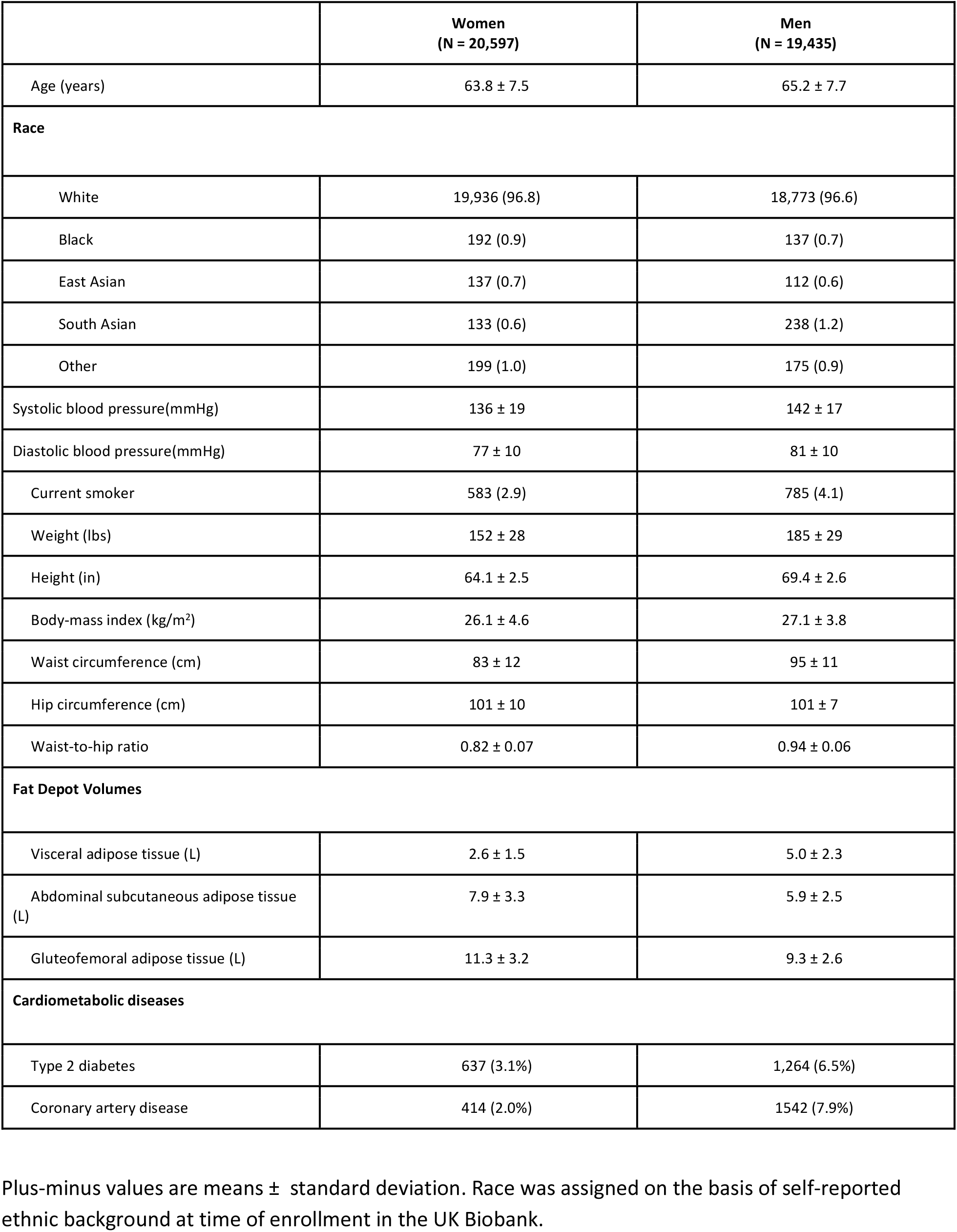
Baseline characteristics of UK Biobank participants at the time of MRI imaging

|  | Women<br>(N = 20,597) | Men<br>(N = 19,435) |
| --- | --- | --- |
| Age (years) | 63.8 ± 7.5 | 65.2 ± 7.7 |
| **Race** |  |  |
| White | 19,936 (96.8) | 18,773 (96.6) |
| Black | 192 (0.9) | 137 (0.7) |
| East Asian | 137 (0.7) | 112 (0.6) |
| South Asian | 133 (0.6) | 238 (1.2) |
| Other | 199 (1.0) | 175 (0.9) |
| Systolic blood pressure(mmHg) | 136 ± 19 | 142 ± 17 |
| Diastolic blood pressure(mmHg) | 77 ± 10 | 81 ± 10 |
| Current smoker | 583 (2.9) | 785 (4.1) |
| Weight (lbs) | 152 ± 28 | 185 ± 29 |
| Height (in) | 64.1 ± 2.5 | 69.4 ± 2.6 |
| Body-mass index (kg/m<sup>2</sup>) | 26.1 ± 4.6 | 27.1 ± 3.8 |
| Waist circumference (cm) | 83 ± 12 | 95 ± 11 |
| Hip circumference (cm) | 101 ± 10 | 101 ± 7 |
| Waist-to-hip ratio | 0.82 ± 0.07 | 0.94 ± 0.06 |
| **Fat Depot Volumes** |  |  |
| Visceral adipose tissue (L) | 2.6 ± 1.5 | 5.0 ± 2.3 |
| Abdominal subcutaneous adipose tissue (L) | 7.9 ± 3.3 | 5.9 ± 2.5 |
| Gluteofemoral adipose tissue (L) | 11.3 ± 3.2 | 9.3 ± 2.6 |
| **Cardiometabolic diseases** |  |  |
| Type 2 diabetes | 637 (3.1%) | 1,264 (6.5%) |
| Coronary artery disease | 414 (2.0%) | 1542 (7.9%) |

Plus-minus values are means ± standard deviation. Race was assigned on the basis of self-reported ethnic background at time of enrollment in the UK Biobank.

### Machine learning facilitates near-perfect estimation of fat depot volumes

We set out to test whether convolutional neural network models could be adequately predictive of VAT, ASAT, and GFAT volumes to enable prediction at scale. We noted that three-dimensional MRI data for 40,032 individuals represented a substantial data burden with almost 58 million axial slices across all participants, corresponding to >18 terabytes of imaging data – a level of complexity that limits computational feasibility for training deep learning models.

To simplify the imaging input into the convolutional neural networks, we transformed three-dimensional MRI images for each participant into two-dimensional coronal and sagittal projections, hypothesizing that this input would prove adequate for highly accurate fat depot volume prediction despite an 830-fold reduction in data input size (**Figure 1**).^31^ Convolutional neural networks – trained on 80% of the participants with fat depots previously quantified – demonstrated near-perfect estimation of each fat depot volume in the 20% of held out individuals (r^2^ = 0.991, 0.991, and 0.978 for VAT, ASAT, and GFAT, respectively) (**Central Illustration; Supplementary Tables S1-S2**). Similar predictive accuracy was noted across male and female participants and across Black, East Asian, and South Asian participants, although sample size was limited in the latter subgroups (**Supplementary Figure S1**). These convolutional neural network models were subsequently applied to the remainder of the 40,032 participants to calculate fat depot volumes.

**FIGURE 1.**
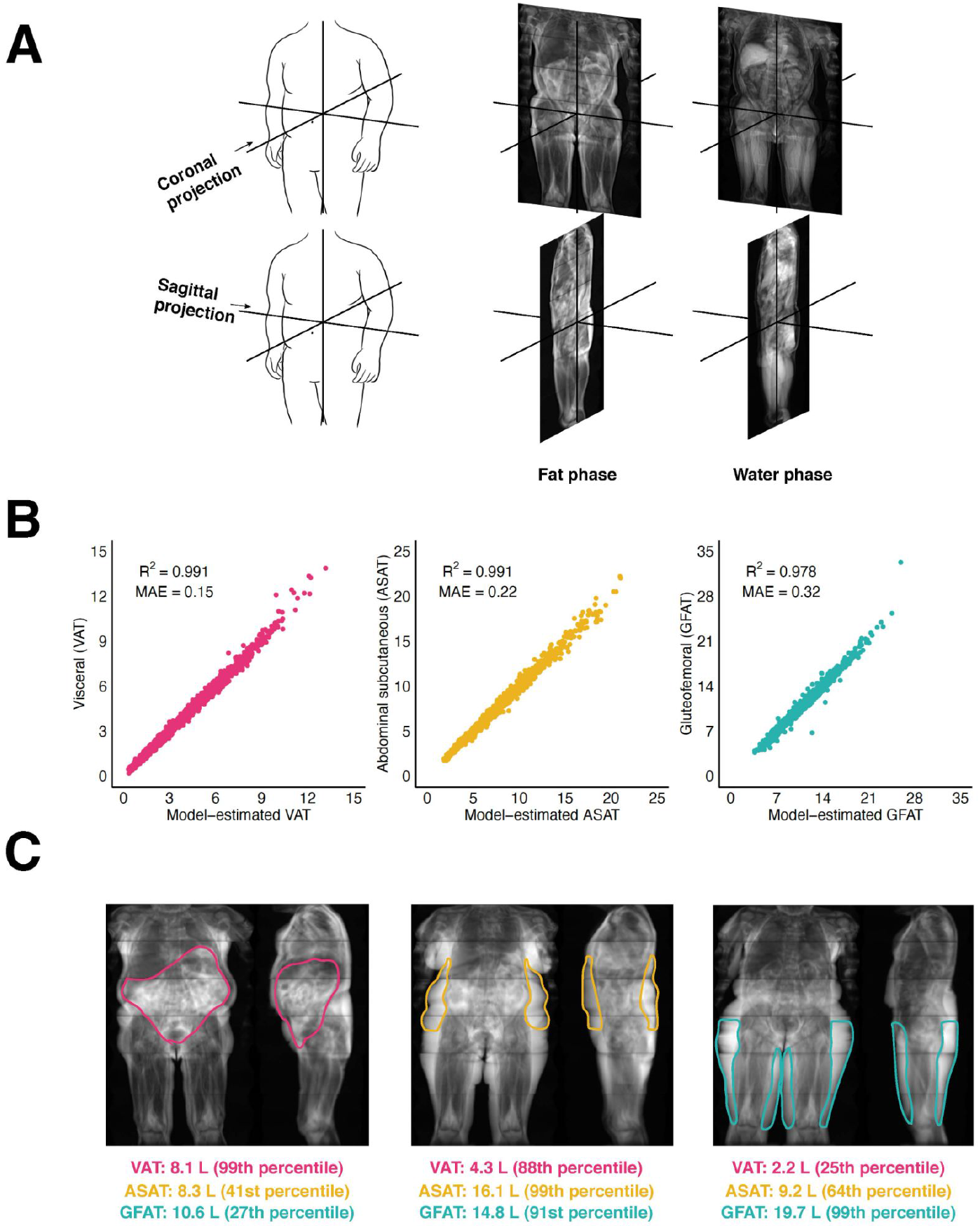
Convolutional neural networks to quantify adipose tissue depots from body MRI images (A) Two-dimensional projections are created by computing the mean pixel intensity along the coronal (top) and sagittal (bottom) axes. Two images were used as inputs into the convolutional neural network: one consisting of the coronal and sagittal two-dimensional projections in the fat phase, and another consisting of the same projections in the water phase. (B) Convolutional neural networks trained on two-dimensional MRI projections achieved near-perfect prediction of each fat depot volume in the holdout set (**Supplementary Table S1**). (C) Three female participants with similar BMI (ranging from 29.1 to 29.6 kg/m^2^) but highly discordant fat depot volumes quantified by convolutional neural networks. Fat depot volume percentiles are computed relative to a subgroup of women with overweight BMI (25 ≤ BMI < 30). Abbreviations: VAT, visceral adipose tissue; ASAT, abdominal subcutaneous adipose tissue; GFAT, gluteofemoral adipose tissue.

### Variation in adipose volumes and association with cardiometabolic diseases

We confirm and extend prior evidence for marked differences in fat depot volume in men versus women (**Figure 2A**).^32,33^ Median visceral adipose tissue volume was substantially higher in men as compared to women – 4.8 versus 2.3 liters, respectively – while subcutaneous and gluteofemoral depots tended to predominate in women (**Table 1**). Moderate strength of correlation BMI and all three fat depots was noted – Pearson *r* ranging from 0.77 to 0.91 – but considerable variation was observed within any clinical BMI category (**Figure 2A-B**).

**FIGURE 2.**
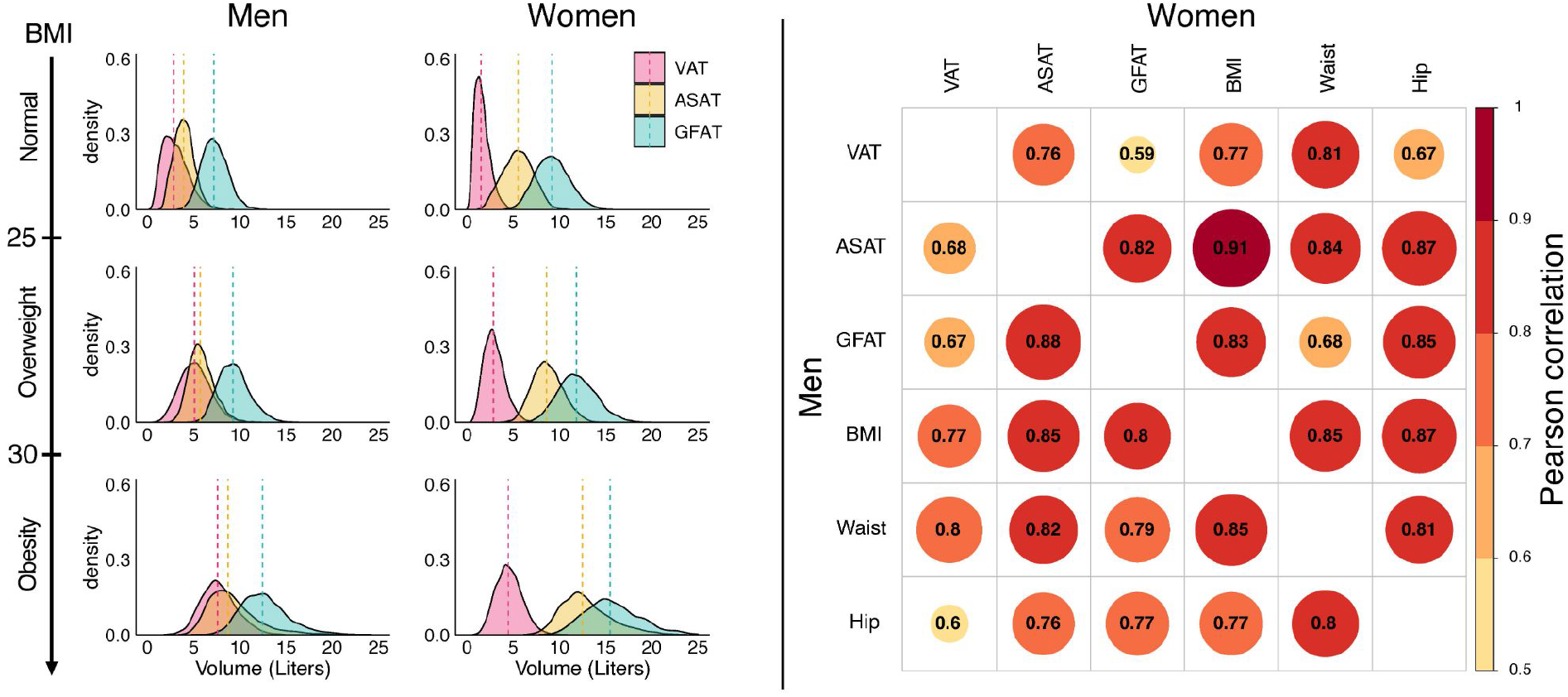
Sex-stratified density plots and correlation plots of visceral, abdominal subcutaneous, and gluteofemoral adipose tissue volumes (A; left) Sex- and BMI-group specific density plots for visceral adipose tissue (VAT), abdominal subcutaneous adipose tissue (ASAT), and gluteofemoral adipose tissue (GFAT). (B; right) Sex-stratified correlation plots between VAT, ASAT, GFAT and three anthropometric measures: body mass index (BMI), waist circumference (Waist), and hip circumference (Hip). Similar plots for BMI-adjusted fat depots are shown in **Supplementary Figures S2-S3**.

Adipose tissue volumes were each associated with increased prevalence of cardiometabolic diseases – as might be expected based on strength of correlation with BMI – with risk gradient somewhat more pronounced for VAT (**Supplementary Table S3-S4**). Taking type 2 diabetes as an example, odds ratios per standard deviation increment (OR/SD) were 2.14 (95%CI 2.05-2.23), 1.69 (95%CI 1.63-1.75), and 1.48 (95%CI 1.42-1.54) for VAT, ASAT, and GFAT, respectively (**Supplementary Table S5**).

### BMI-adjusted local fat depots and cardiometabolic disease

To disentangle the unique impact of each fat depot from overall BMI, we next generated new measurements of VATadjBMI, ASATadjBMI, and GFATadjBMI for each participant by computing sex-specific BMI residuals in 38,680 (97%) of the study population with BMI measurement on the day of MRI imaging available. These residuals reflect the difference in an individual’s fat depot volume as compared with that expected based on BMI. These metrics were fully independent of BMI and largely independent of anthropometric measures and each other (**Supplementary Figures S2-S3**).

In contrast to analysis of raw tissue volumes – where each depot was associated with increased risk – significant heterogeneity was noted for BMI-adjusted values. In a mutually adjusted logistic regression model, we observe that VATadjBMI was associated with increased prevalence of type 2 diabetes – OR/SD 1.49; 95% CI 1.43-1.55). By contrast, a largely neutral effect estimate was noted for ASATadjBMI (OR/SD 1.08; 95%CI 1.03-1.14) and GFATadjBMI volumes were associated with decreased risk (OR/SD 0.75; 95% CI: 0.71-0.79) (**Figure 3**). Effect estimates were largely consistent in subgroups binned by age or sex, with somewhat amplified associations in participants with BMI less than 25 (**Supplementary Figure S4-S5**). A similar pattern was observed for coronary artery disease, where associations for VATadjBMI, ASATadjBMI, and GFATadjBMI were 1.17 (95%CI 1.11-1.22), 1.00 (95%CI 0.94-1.05), and 0.89 (95%CI 0.84-0.94), respectively.

**FIGURE 3.**
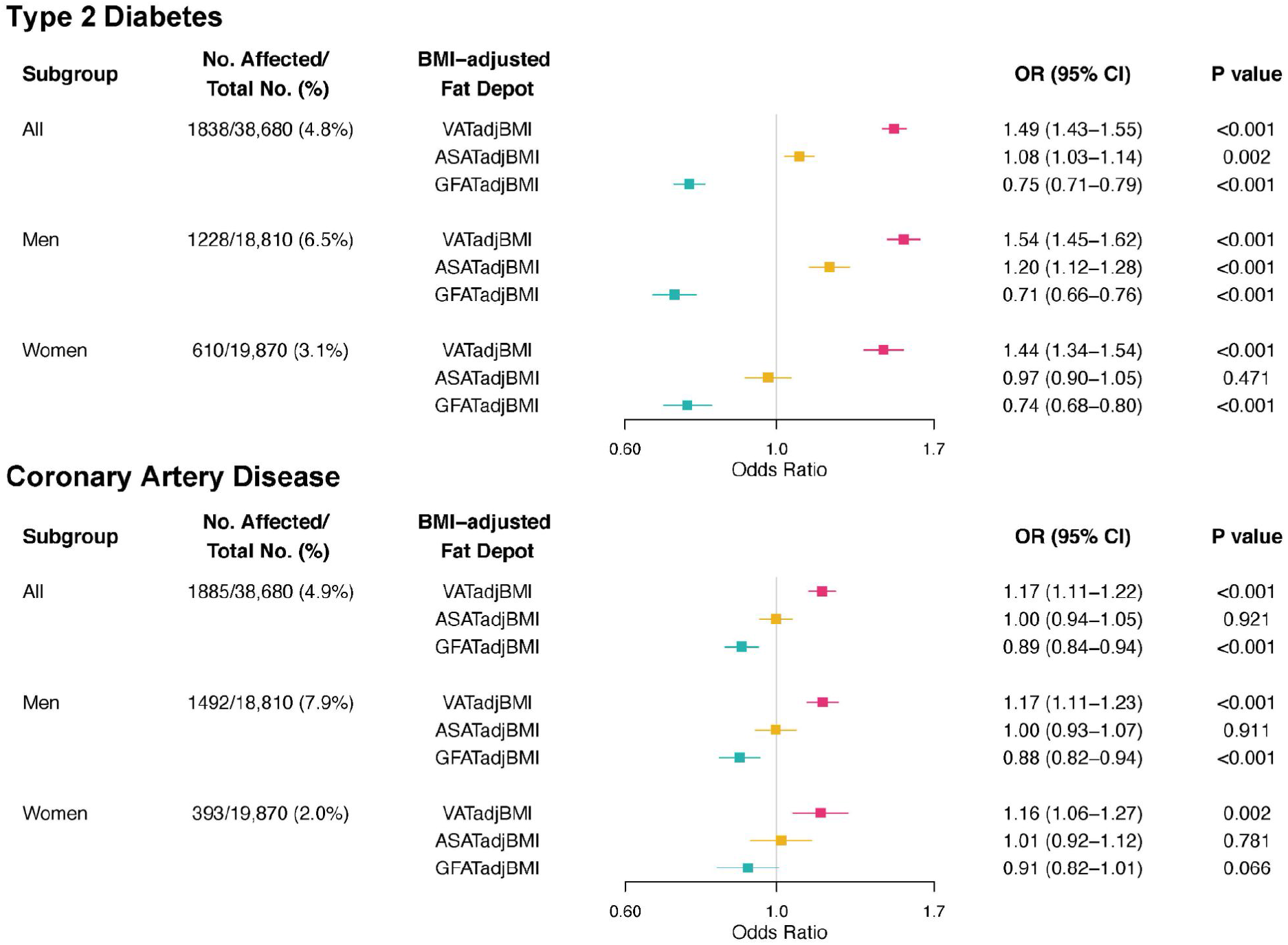
Association of body-mass index adjusted fat depots with type 2 diabetes and coronary artery disease (A; top) Odds ratios per standard deviation shown for prevalent type 2 diabetes. (B; bottom) Odds ratios per standard deviation shown for prevalent coronary artery disease. Logistic regression models were adjusted for age, sex (except in sex subgroup analyses), BMI, the other two fat depots (e.g. ASATadjBMI and GFATadjBMI for VATadjBMI), and MRI imaging center.

To better understand the gradients in absolute prevalence rates according to BMI-adjusted fat depots, we calculated standardized estimates for the lowest quintile, quintiles 2-4, and the highest quintile within clinical categories of normal, overweight, and obese participants.

Using this approach, we note substantial gradients in the prevalence of cardiometabolic diseases according to local adipose tissue burden, even within clinical BMI categories (**Figure 4, Supplementary Table S6-S7**). As a representative example, men with normal BMI but VATadjBMI in the highest quintile had a probability of type 2 diabetes of 6.6% (95%CI 5.5-7.9), higher than obese men with VATadjBMI in the lowest quintile, in whom probability was 5.2% (95%CI 4.1-6.6). Among obese women, estimates of diabetes ranged from 3.5 to 9.2% across quintiles of VATadjBMI but 7.6 to 3.6% for GFATadjBMI. A similar pattern – with less pronounced gradients – was observed for coronary artery disease.

**FIGURE 4.**
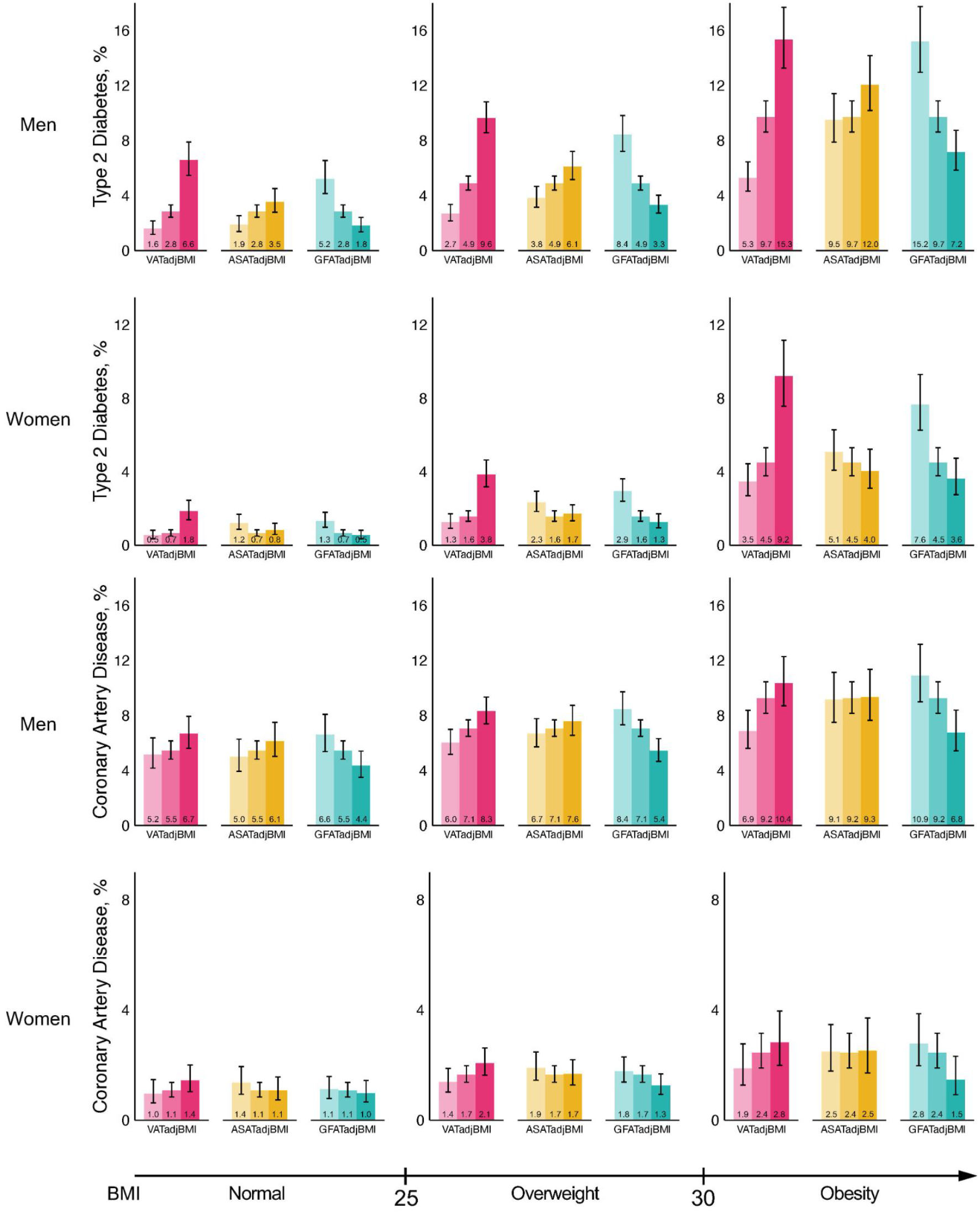
Prevalence of type 2 diabetes and coronary artery disease, according to quintiles of body-mass index adjusted fat depot and body-mass index strata For each fat depot, the three bars from lightest to darkest represent the bottom quintile, quintiles 2-4, and the top quintile of the BMI-adjusted fat depot in question, respectively. Mean body-mass index was 26.5 kg/m^2^ with 15,446 (39.9%) individuals with BMI < 25, 16,179 (41.8%) with 25 ≤ BMI < 30, and 7055 (18.2%) with BMI ≥ 30.

### BMI-adjusted fat depots and risk of incident cardiometabolic diseases

Over a median follow-up of 2.8 years, 235 (0.6%) and 607 (1.6%) participants had a new diagnosis of type 2 diabetes or coronary artery disease recorded in the electronic health record. BMI-adjusted fat depots were similarly associated with risk of future disease events in mutually adjusted models. For incident type 2 diabetes, hazard ratios per SD increase (HR/SD) were 1.45 (95%CI 1.30-1.61), 0.96 (95%CI 0.84-1.08), and 0.84 (95%CI 0.74-0.95) for VATadjBMI, ASATadjBMI, and GFATadjBMI respectively (**Supplementary Table S8**). For incident coronary artery disease, HR/SD) were 1.17 (95%CI 1.08-1.26), 1.04 (95%CI 0.95-1.14), and 0.91 (95%CI 0.83-1.00) for VATadjBMI, ASATadjBMI, and GFATadjBMI respectively.

## DISCUSSION

In this study, we demonstrated that a deep learning approach based on two-dimensional MRI projections is adequately predictive to quantify VAT, ASAT, and GFAT volumes at scale. By then moving away from raw fat depot volumes – which are driven largely by BMI and overall adiposity – to BMI-adjusted measurements, we demonstrated a consistent trend of VATadjBMI associated with increasing risk of type 2 diabetes and coronary artery disease, ASATadjBMI largely risk-neutral, and GFATadjBMI conferring protection. These results have at least four implications.

First, machine learning can enable new insights from large-scale data repositories of difficult-to-measure phenotypes. In this study, convolutional neural network models were used to precisely measure fat depot measurements from MRI images, considered the gold standard modality for volumetric measurement of adipose tissue.^16,34^ Hypothesis-informed simplification of the input data – in this study moving from three-dimensional MRI images to two-dimensional MRI projections – was necessary to ensure computational feasibility. This work adds to several recent studies of machine learning-derived phenotypes including aortic size, liver fat, and cardiac trabecular structure.^35–37^ Although population-based assessment of fat distribution using MRI is unlikely to be practical, these results lay the scientific foundation for efforts to quantify such measures using other data – such as DEXA images or abdominal CT scans already embedded in the electronic medical record for some patients – or even static images of body silhouette, as might conceivably be obtained with a smartphone camera.^38,39^

Second, these results support a growing appreciation that various fat depots – rather than serving as an agnostic sink for energy storage – have distinct metabolic profiles. Previous work has noted significant functional differences in adipocytes according to specific fat depot, ascribed in part to site-specific expression of developmental genes associated with adipogenesis.^40,41^ While VAT tends to be the primary site for immediate storage of dietary-derived fat via adipocyte hypertrophy and has a higher rate of lipid turnover, GFAT is a more stable fat depot that primarily expands via adipocyte hyperplasia and may spare expansion of harmful visceral or ectopic fat depots. These and other studies support a natural order of fat deposition, whereby a primary driver of high VAT in specific individuals may reflect an inability to adequately expand ASAT or GFAT depots.^13,42^ In rare Mendelian lipodystrophies – as occurs in individuals who harbor pathogenic *LMNA* mutations – an extreme example of this paradigm leads to marked reduction of ASAT and GFAT but increased VAT and increased rates of severe insulin resistance.^43^ Whether individuals in the extreme tails of low GFATadjBMI and ASATadjBMI or high VATadjBMI might be enriched for genetic perturbations in lipodystrophy genes or the inherited component to these metrics is largely ‘polygenic’ – due to the aggregate effects of many common DNA variants, each of modest effect size – warrants further study.^44,45^

Third, changes in measures of local adiposity – independent of weight and body-mass index – may serve as reliable proxies of cardiometabolic benefits of a given intervention, and warrant consideration as additional endpoints for future clinical trials. Most studies to date of obesity interventions have focused on reduction in overall weight or BMI as the primary outcome, consistent with FDA regulatory guidance.^45^ However, at least two classes of drugs appear to have a selective VAT reduction effect in clinical trials: thiazolidinediones and a synthetic form of growth hormone releasing hormone.^46,47^

Whether these therapies might be repurposed from their original indications – type 2 diabetes and HIV-associated lipodystrophy – or new agents might prove useful in a subset of individuals with VAT-driven increases in cardiometabolic risk warrants further study.

Fourth, although our data suggests similar performance of the deep learning models across sex and race subgroups, additional validation across ancestrally and geographically diverse populations would be of considerable value, especially given prior evidence of significant variability in fat distribution indices across racial groups.^46,47^ An important example relates to the South Asian population, where abnormal fat distribution has been postulated as a key driver of the markedly increased rates of cardiovascular disease and diabetes observed, often in the context of a relatively normal BMI.^48,49^

## STUDY LIMITATIONS

Our study has several limitations. First, this study was a cross-sectional analysis of individuals with a mean age of 65 years at time of imaging. Future studies of individuals across the lifespan – especially those that include repeat imaging assessments – are warranted. Second, although we note striking associations of BMI-adjusted fat depots with cardiometabolic disease, these observational data do not definitely prove causation or that modification of fat distribution will lead to therapeutic gain.

## CONCLUSION

In conclusion, we used a machine learning approach based on two-dimensional projections of body MRI data to compute VAT, ASAT, and GFAT volumes at scale in 40,032 individuals of the UK Biobank. BMI-adjusted fat depot measurements displayed divergent associations with cardiometabolic diseases and were shown to alter risk within BMI subgroups. These BMI-adjusted metrics may serve as useful additional endpoints for obesity interventions to more completely capture metabolic health associated with body composition.

## Data Availability

The raw UK Biobank data - including the anthropometric data reported here - are made available to researchers from universities and other research institutions with genuine research inquiries following IRB and UK Biobank approval. Code used to ingest whole-body Dixon MRI images from UK Biobank participants is made available at the following Github repository under an open-source BSD license: https://github.com/broadinstitute/ml4h/tree/master/ml4h/applications/ingest.

## ACKNOWLEDGEMENTS

The authors thank Mary O’Reilly of the Broad Institute’s Pattern data visualization team for assistance in graphic and visual design.

