## Supplementary Appendix for "Association of machine learning-derived measures of body fat distribution with cardiometabolic diseases in >40,000 individuals"

**Table of Contents**

**Supplementary Methods**

**Figure S1** Convolutional neural network performance among sex and race subgroups

**Figure S2** Density plots of VATadjBMI, ASATadjBMI, and GFATadjBMI

**Figure S3** Correlogram of fat depots adjusted-for-BMI and anthropometric measurements

**Figure S4** Fat depot specific effects on type 2 diabetes across age, sex, and BMI subgroups

**Figure S5** Fat depot specific effects on coronary artery disease across age, sex, and BMI subgroups

**Table S1** Characteristics of 80% training and 20% testing sets for fat depot volume CNNs

**Table S2** Convolutional neural network performance for adipose tissue volumes

**Table S3** Type 2 Diabetes definition

**Table S4** Coronary artery disease definition

**Table S5** Cardiometabolic disease associations with BMI-unadjusted visceral, abdominal subcutaneous, and gluteofemoral adipose tissue volumes

**Table S6** Standardized prevalence of type 2 diabetes across quintiles of VATadjBMI, ASATadjBMI, and GFATadjBMI

**Table S7** Standardized prevalence of coronary artery disease across quintiles of VATadjBMI, ASATadjBMI, and GFATadjBMI

**Table S8** Hazard ratios of VATadjBMI, ASATadjBMI, and GFATadjBMI for incident cardiometabolic disease

### Supplementary Methods

#### Raw MRI imaging data in the UK Biobank

We studied body magnetic resonance imaging (MRI) data from 43,531 participants of the UK BIobank study. As previously described, the UK Biobank is an observational study that enrolled over 500,000 between the ages of 40 and 69 years between 2006 and 2010.[1] In an ongoing effort, up to 100,000 participants from four regions in the UK will be asked to return for comprehensive imaging – at the time of this study, 43,531 participants had been imaged and had MRI imaging data available for download.[2]

Participants in the MRI imaging substudy were scanned using Siemens Aera 1.5-T MRI scanners from the neck to the knees using the Dixon method, an MRI sequence that can be used to isolate fat from water signal.[3] Image acquisition from the neck to the knees was comprised of 6 contiguous stages with varying numbers of axial slices: stage 1 covered the neck with 64 slices, stages 2-4 covered the torso with 44 slices each, stage 5 covered the upper legs with 72 slices, and stage 6 covered the lower legs with 64 slices.[2]

The output of this procedure is a set of four MRI sequences for each participant: in-phase and out-of-phase ("phase" in reference to water and fat molecule precessions), and fat-only and water-only (obtained by adding and subtracting in-phase and out-of-phase, respectively).

#### Fat depots previously quantified in a subset of UK Biobank participants

Among UK Biobank participants who underwent MRI imaging study, a subset had visceral adipose tissue (VAT) volume, abdominal subcutaneous adipose tissue (ASAT) volume, and total adipose tissue between the bottom of the thigh muscles to the top of vertebrae T9 (TAT) volume quantified and made available via the UK Biobank portal to the broader research community.[4–9] VAT (field 22407, "volume of the adipose tissue within the abdominal cavity, excluding adipose tissue outside the abdominal skeletal muscles and adipose tissue and lipids within and posterior of the spine and posterior of the back muscles") was available in 9,978 participants, ASAT (field 22408, "volume of the subcutaneous adipose tissue in the abdomen from the top of the femoral head to the top of the thoracic vertebrae T9") was available in 9,979, and TAT (field 22415, "total volume of adipose tissue, measured by MRI, between the bottom of the thigh muscles to the top of vertebrae T9") was available in 8,524. Based on these definitions, we additionally computed gluteofemoral adipose tissue (GFAT) volume:

GFAT = TAT (between top of T9 and bottom of thigh muscles) - VAT - ASAT

Given that the vast majority of adipose tissue between the top of vertebrae T9 and the top of the femoral head is accounted for by VAT or ASAT, GFAT was defined as total adipose tissue between the top of the femoral head and the bottom of the thigh muscles.

### Simplifying three-dimensional MRI images to two-dimensional mean projections

Each of the 6 contiguous stages that make up an individual's MRI in the UK Biobank are acquired at slightly different resolutions, and so a pre-processing step was required prior to subsequent analysis. Resolutions ranged from 2.232 x 2.232 x 4.5 $mm^3$ (stages 2-4) to 2.232 x 2.232 x 3.0 $mm^3$ (stage 1). We resampled each series to the highest available resolution (voxel = 2.232 x 2.232 x 3.0 $mm^3$), enabling a merged single three-dimensional volume that included all stages.

We next evaluated the computational burden associated with training machine learning models on 3D MRI data. We noted that three-dimensional MRI data for >40,000 individuals represented a substantial data burden with almost 58 million axial slices across all participants, corresponding to >18 terabytes of imaging data – a level of complexity that limits computational feasibility for training deep learning models.

To simplify the machine learning model inputs, we transformed MRI images for each participant into two-dimensional projections of the coronal and sagittal anatomical planes – hypothesizing that this input would prove adequate for accurate fat depot volume quantification. Similar reduced representations of MRI imaging data were used in a recent study with excellent model performance.[10] Coronal and sagittal two-dimensional projections were generated by computing the mean intensity projection in each orientation. For example, a given pixel on a coronal two-dimensional projection represents the mean intensity across all pixels making up a line oriented in the anterior-posterior direction perpendicular to the coronal plane. This procedure was done for each MRI sequence for each participant.

### Quality control of MRI imaging data

Starting with raw imaging data for 43,531 participants, 389 participants had either incomplete acquisitions (e.g. missing a stage), or faulty or corrupted data (e.g. length of pixel data does not match the metadata records). After exclusion of these 389 participants, we performed quality control of the remaining 43,142 sets of two-dimensional projections and noted 7 classes of imaging artifacts that occurred:

- Fat/water swaps involving a complete stage, N = 412
- Knee(s) missing and/or metal artifacts in the body, N = 257
- Field-of-view errors caused by a misaligned individual in either superior-inferior direction such that the head or chin was partly or fully visible or the clavicle was not fully visible, N = 397
- Participant was too large to be completely captured in the scanner view, N = 107
- Instance swaps with self-contained areas of fat/water swaps within a stage, N = 325
- Liver swaps where fat/water instance swaps are restricted to the superior part of the liver, N = 1,289
- Technically unusable because of unexpected observations such as stage-wide duplications or stages in the incorrect order, blank stages, or extremely noisy acquisitions, N = 323

In total, 3,499 (8.0%) of individuals were excluded by our quality control pipeline – consistent with a prior estimate of imaging artifact in UK Biobank body MRI data – resulting in 40,032 participants who were carried forward into downstream analysis.[6]

**Convolutional neural networks (CNNs) to quantify fat depots**

Among the 40,032 individuals who remained after quality control, 9,040 participants had VAT quantified, 9,041 participants had ASAT quantified, and 7,754 participants had GFAT quantified and made available via previous studies.[4–9] For each fat depot volume, participants were split into 80% for model training and 20% for model validation (**Table S1**). We used 5-fold cross-validation within the 80% model training data to estimate error.

For each of these three fat depot volumes, a CNN was trained on a pair of fat phase and water phase images, where each image was composed of (a) a coronal two-dimensional projection and (b) a sagittal two-dimensional projection of the body MRI to predict each fat depot volume. Each CNN was developed with the DenseNet-121 architecture pre-trained on ImageNet as the base model.[11,12] The last dense block output was flattened using a global average pooling layer and then fed into three fully connected layers of size 64, 256, and 1, with the last layer having no activation function (linear mapping). All other activation functions use the ReLU non-linearity. All models were trained using the Adam optimizer with a learning rate set to a cosine decay policy decaying from 0.001 to 0 over 100 epochs, a shrinkage loss function using the hyperparameters a = 10.0 and c = 0.2, and a batch size of 32.[13,14]

For all training data, the following augmentations (random permutations of the training images) were applied: random shifts in the XY-plane by up to ±16 pixels, rotations by up to ±5 degrees around its center axis, and the coronal view horizontally flipped with a probability of 50%. Each view (coronal and sagittal) were separately pre-normalized by its z-score (0 mean, standard deviation of 1), followed by joint normalization following concatenation side-by-side.

Performance of each CNN developed here is shown in **Table S2**.

**Code availability**

Code used to ingest whole-body Dixon MRI images from UK Biobank participants is made available at https://github.com/broadinstitute/ml4h/tree/master/ml4h/applications/ingest under an open-source BSD license.

**Figure S1 Convolutional neural network performance among non-White racial groups**

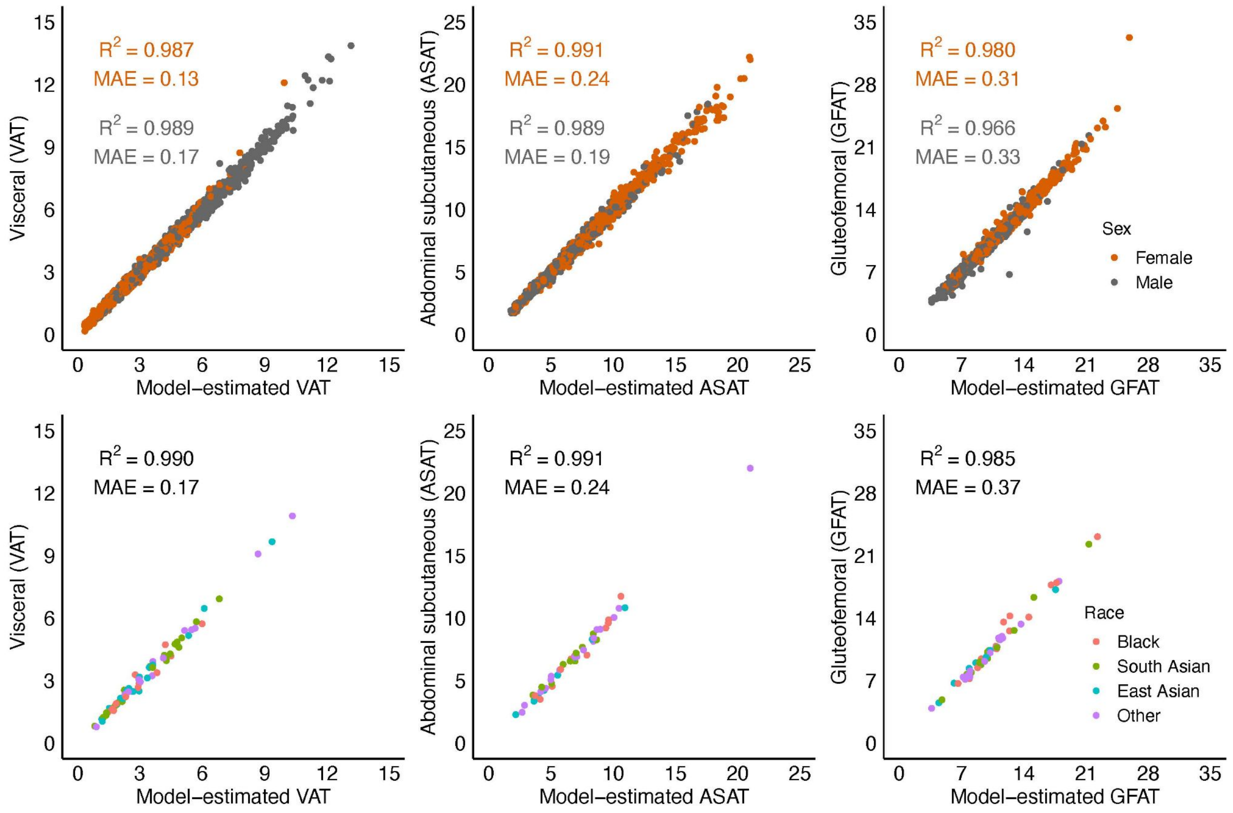


Volumes in Liters are shown on both axes. (top) Performance stratified by sex. (bottom) Performance stratified by self-identified racial group; 63 of 1,804 individuals in the VAT holdout set, 48 of 1,815 individuals in the ASAT holdout set, and 52 of 1,551 individuals in the GFAT holdout set were non-White in the holdout set.

**Figure S2 Density plots of VATadjBMI, ASATadjBMI, and GFATadjBMI**

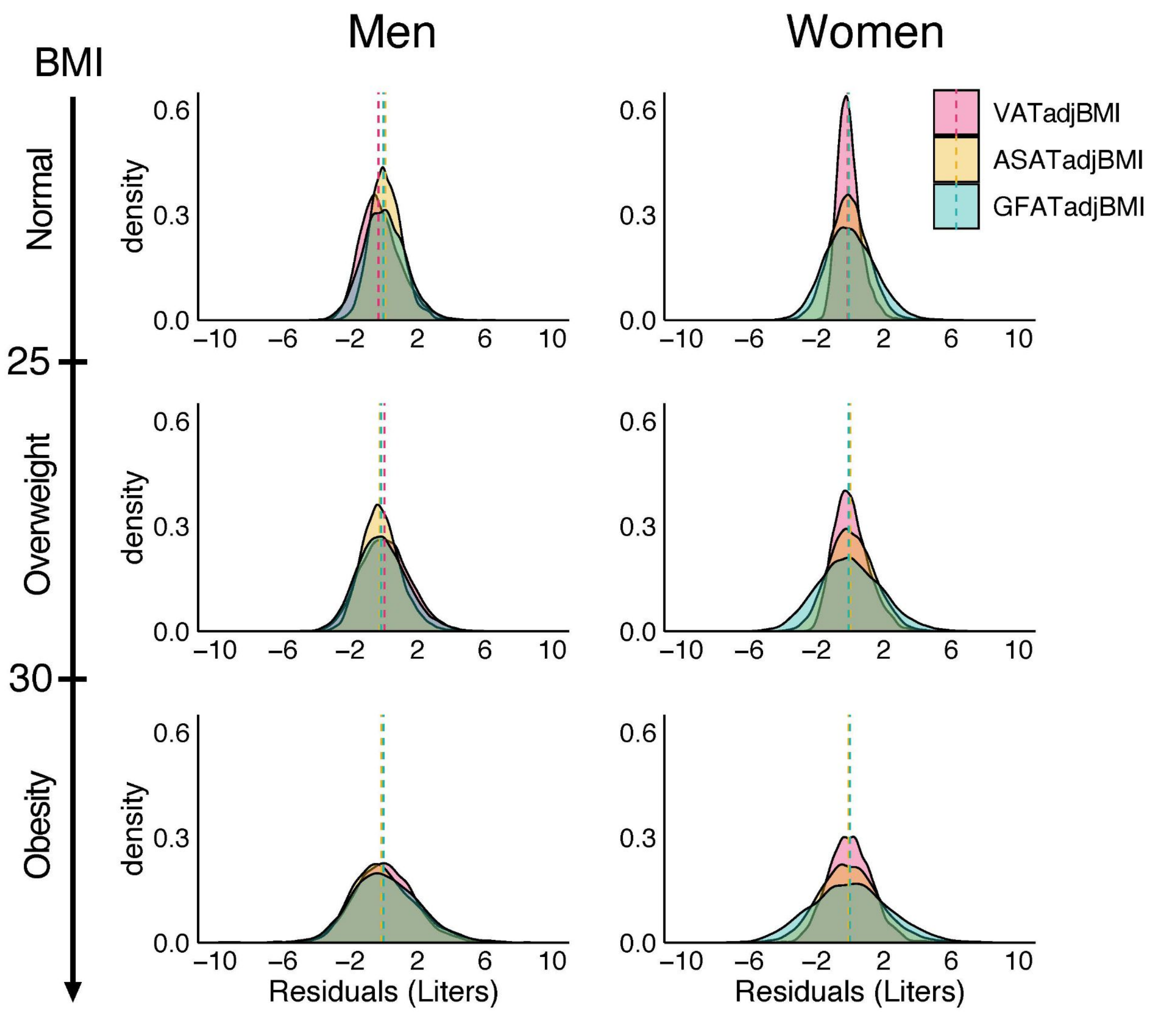


Sex- and BMI-group specific density plots for visceral adipose tissue volume adjusted for body-mass index (BMI), (VATadjBMI), abdominal subcutaneous adipose tissue volume adjusted for BMI (ASATadjBMI), and gluteofemoral adipose tissue volume adjusted for BMI (GFATadjBMI).

**Figure S3 Correlogram of fat depots adjusted-for-BMI and anthropometric measurements**

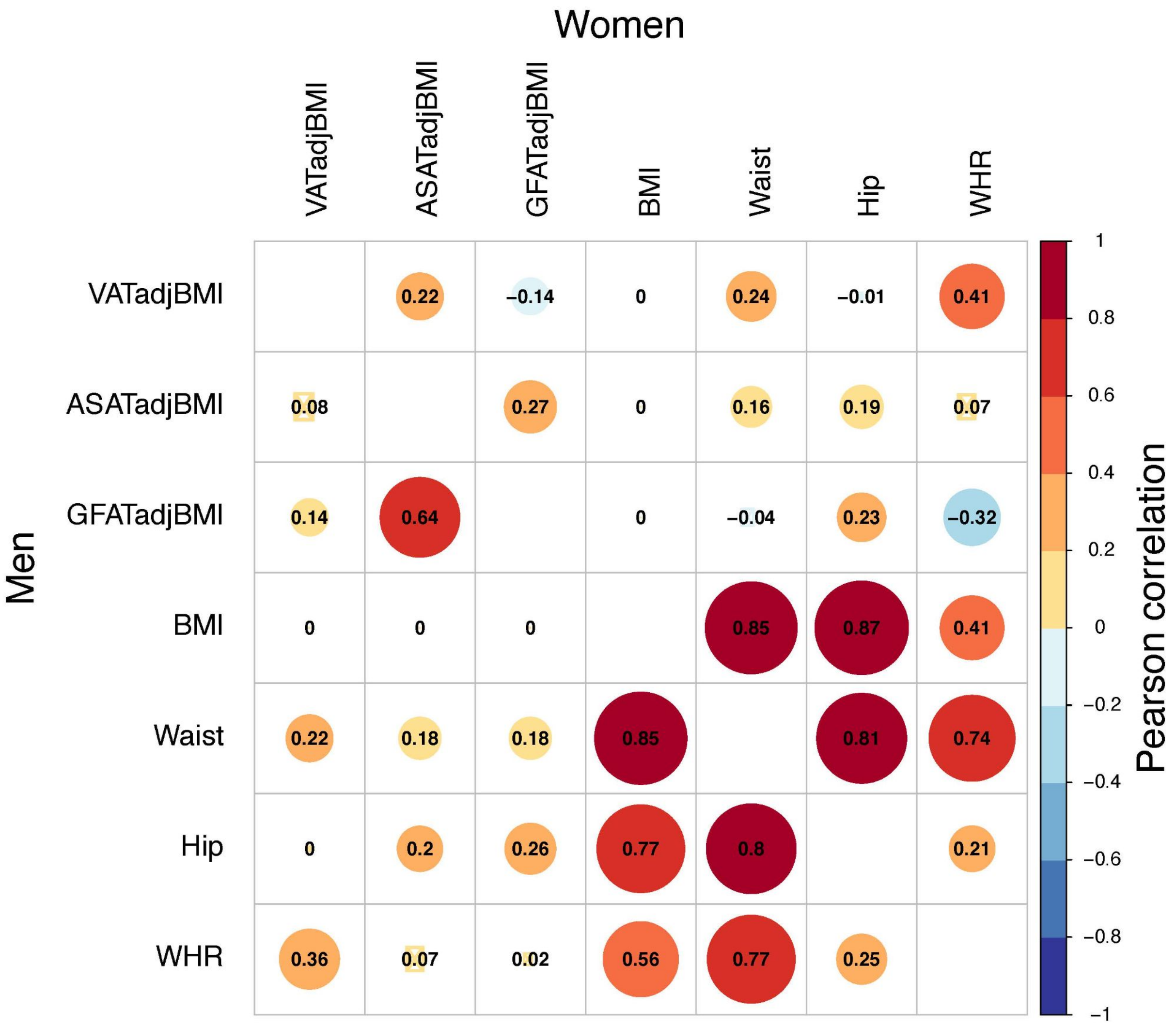


Sex-stratified correlation plots between visceral adipose tissue volume adjusted for BMI (VATadjBMI), abdominal subcutaneous adipose tissue adjusted for BMI (ASATadjBMI), gluteofemoral adipose tissue volume adjusted for BMI (GFATadjBMI) and four anthropometric measures: body mass index (BMI), waist circumference (Waist), hip circumference (Hip), and waist-hip ratio (WHR).

**Figure S4 Fat depot specific effects on type 2 diabetes across age, sex, and BMI subgroups**

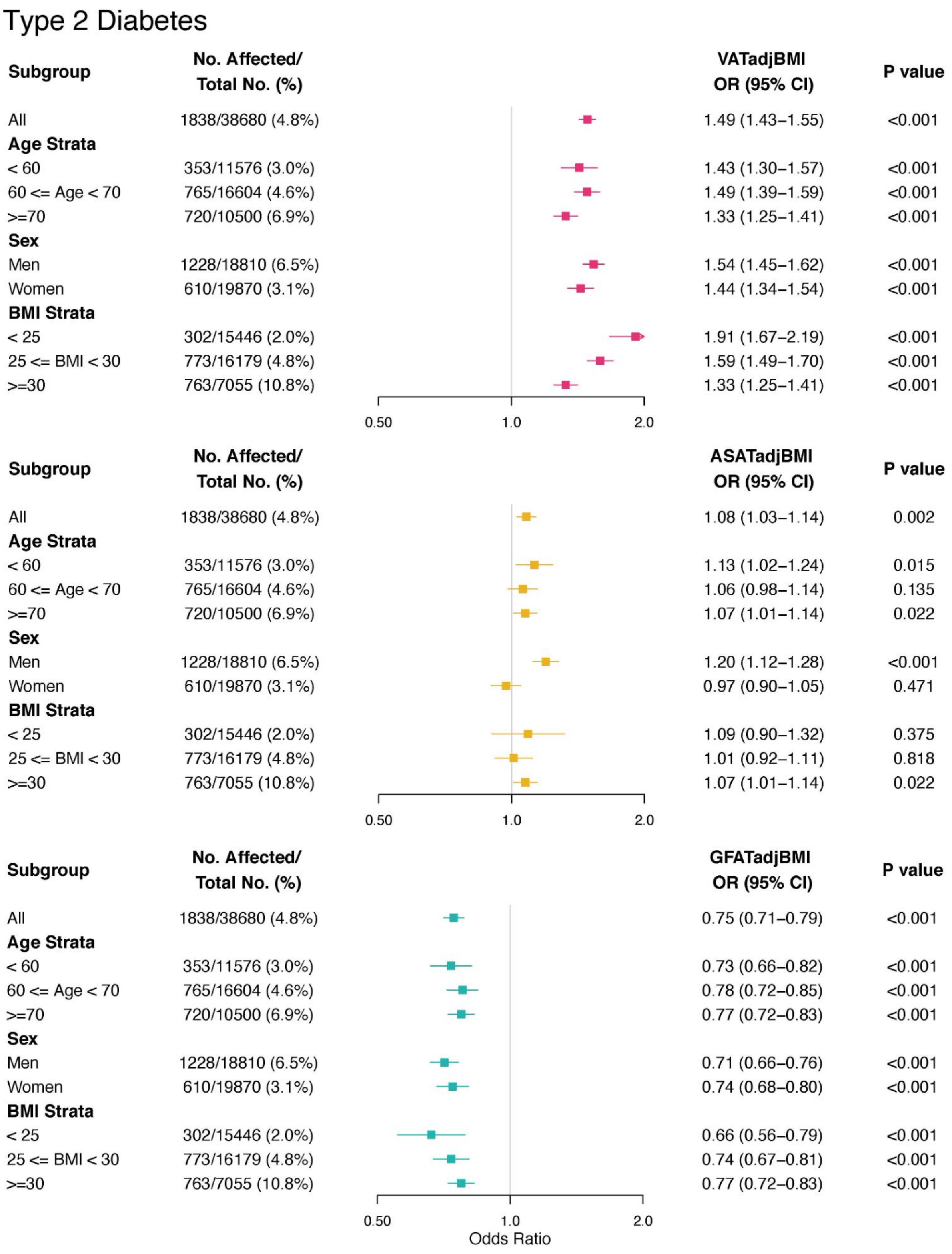

**Figure S5 Fat depot specific effects on coronary artery disease across age, sex, and BMI subgroups**

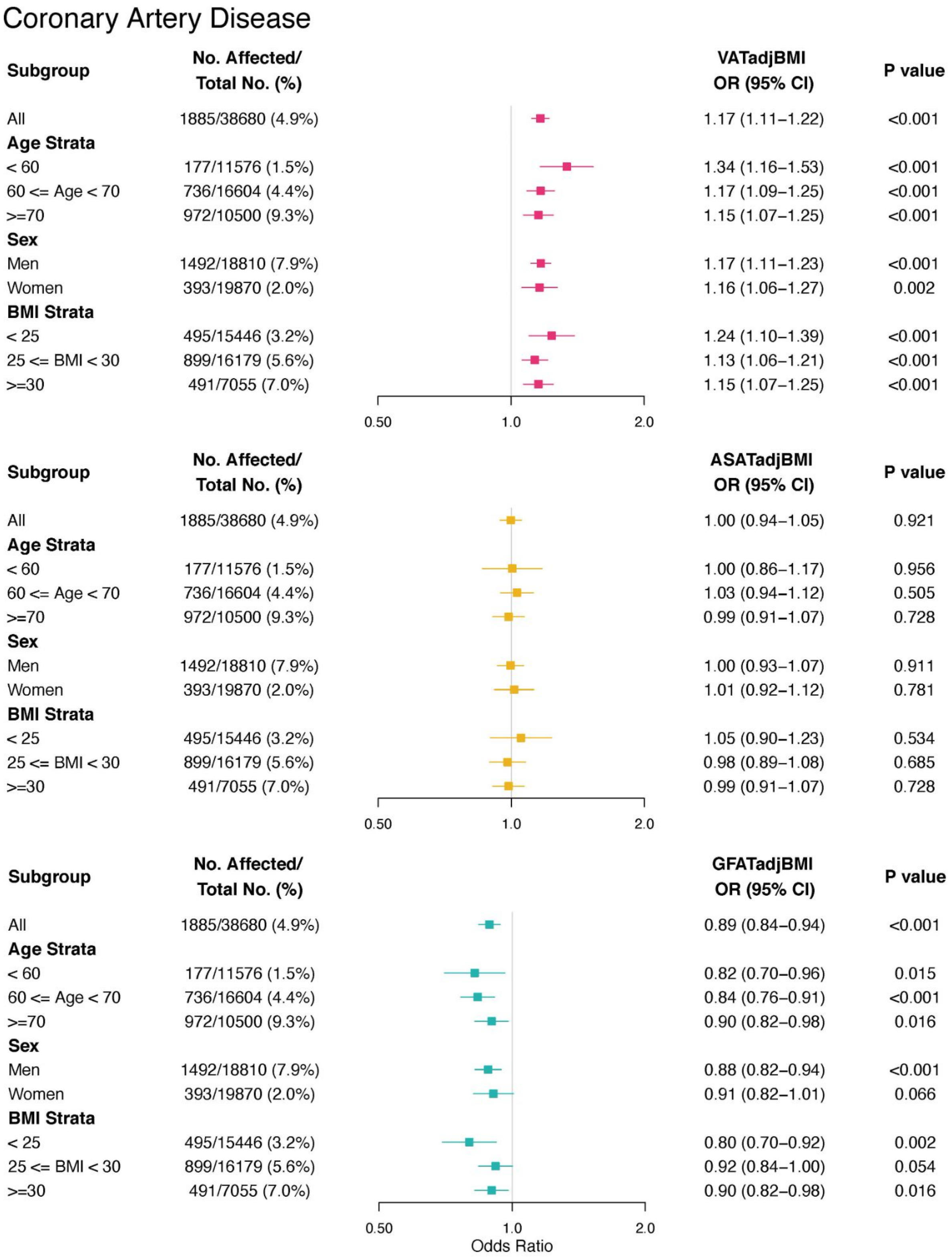

**Table S1 Characteristics of 80% training and 20% testing sets for fat depot volume CNNs**

| | **Visceral adipose tissue** | | **Abdominal subcutaneous adipose tissue** | | **Gluteofemoral adipose tissue** | |
|---|---|---|---|---|---|---|
| | **Development** | **Holdout** | **Development** | **Holdout** | **Development** | **Holdout** |
| **Count (N)** | 7,236 | 1,804 | 7,226 | 1,815 | 6,203 | 1,551 |
| **Age (years)** | 62.6±7.5 | 62.9±7.4 | 62.6±7.5 | 62.8±7.4 | 62.7±7.5 | 62.3±7.6 |
| **# Men (%)** | 3,505 (48.4) | 888 (49.2) | 3,507 (48.5) | 887 (48.9) | 2,891 (46.6) | 702 (45.3) |
| **Weight (lb)** | 169.4±32.9 | 167.7±32.4 | 168.8±32.6 | 170.2±33.5 | 166.9±31.0 | 167.4±31.7 |
| **Height (in)** | 66.7±3.6 | 66.6±3.7 | 66.7±3.6 | 66.7±3.6 | 66.5±3.6 | 66.5±3.5 |
| **BMI (kg/m$^2$)** | 26.8±4.3 | 26.6±4.2 | 26.7±4.3 | 26.9±4.5 | 26.6±4.2 | 26.7±4.3 |
| **Waist circumference (cm)** | 87.8±12.1 | 87.5±12.0 | 87.6±12.0 | 88.1±12.4 | 87.1±11.7 | 87.3±11.9 |
| **Hip circumference (cm)** | 101.6±8.5 | 101.3±8.3 | 101.5±8.3 | 101.7±8.8 | 101.2± 8.2 | 101.3±8.5 |
| **Waist-hip ratio** | 0.86±0.08 | 0.86±0.08 | 0.86±0.08 | 0.86±0.08 | 0.86±0.08 | 0.86±0.08 |
| **Fat depot volume (L)** | 3.8±2.2 | 3.7±2.2 | 7.1±3.1 | 7.1±3.2 | 10.4±3.1 | 10.5±3.2 |

Participants were randomly split into 80/20% development/holdout datasets for each fat depot. All values are reported as mean ± std unless otherwise specified. Across all listed variables, no differences between development and holdout sets across the three phenotypes were significant at the $p < 0.05$ level.

**Table S2 Convolutional neural network performance for adipose tissue volumes**

| | Previously quantified volumes | | | Deep learning performance | |
|---|---|---|---|---|---|
| **Phenotype** | **N** | **[min, max]** | **mean ± std** | **MAE (mean ± std)** | **$R^2$ (mean ± std)** |
| Visceral adipose tissue (VAT) volume | 9,040 | [0.15, 14.41] | 3.77 ± 2.23 | 0.151 ± 0.0010 | 0.991 ± 0.0004 |
| Abdominal subcutaneous adipose tissue (ASAT) volume | 9,041 | [1.43, 23.48] | 7.07 ± 3.15 | 0.221 ± 0.0019 | 0.991 ± 0.0001 |
| Gluteofemoral adipose tissue (GFAT) volume | 7,754 | [3.36, 33.27] | 10.39 ± 3.13 | 0.321 ± 0.0042 | 0.975 ± 0.006 |

Performance was assessed using evaluation on data unseen to the models during training (holdout) with 5-fold cross-validation to estimate errors. Abbreviations: MAE, mean absolute error; std, standard deviation.

**Table S3 Type 2 diabetes definition.**

| Description | Fields | Description |
|---|---|---|
| **Step 1**: Select individuals with self-reported type 2 diabetes in nurse interview, relevant inpatient ICD-10 code, or relevant death registry record. | 20002 (self-report), 41202/41204 (ICD-10), 40001/40002 (death register) | 20002: 1223<br>41202/41204: E11, E11.0 - E11.9<br>40001/40002: E11, E11.0 - E11.9 |
| **Step 2A**: Identify individuals with a single recorded Hemoglobin A1C >= 6.5% on the date of or prior to date of imaging (Instance 2) | 30750 | Select converted value >= 6.5% |
| **Step 2B**: Identify individuals who report taking non-insulin diabetes medication and self-report history of diabetes | 20002 (medical history),<br>20003 (medication history) | 20002: 1220, 1221, 1223<br>20003: 1140884600,1140857584,1140874706,1140874664,1140874674,1140874718,1140857494,1140874744,1140874646,1140874658,1141152590,1140874712,1140874666,1140874650,1140874724,1140874726,1140874728,1140874736,1140910564,1140874732,1140874652,1141169504,1140857496,1140874746,1140868908,1140857508,1141157284,1140874660,1141177606,1141189094,1141189090,1141177600,1140874686,1141171646,1141168660,1141173882,1141173786,1141153254,1141171652,1140882964,1140868902 |
| **Step 2C**: Identify individuals who are using insulin without a history of type 1 diabetes | 20002 (medical history),<br>6153/6177 (insulin history) | 20002: 1222<br>6153/6177: value = 3 |
| **Step 3**: Define prevalent type 2 diabetes as any individual who satisfies Step 1 with recorded date before date of imaging OR satisfies Step 2A-C | | |
| **Step 4**: Define incident type 2 diabetes as any individual who satisfies Step 1 with a recorded date after the date of imaging | | |

**Table S4 Coronary artery disease definition.**

| **Fields** | **Codes** |
|---|---|
| 20002 (self-report medical history) | 1075 |
| 20004 (self-report surgical history) | 1070, 1095, 1523 |
| 6150 (doctor diagnosed diseases) | 1 |
| 41202/41204 (ICD-10 codes), 40001/40002 (death registry) | I21, I21.0-I21.4, I21.9, I22, I22.0-I22.1, I22.8-I22.9, I23, I23.0-I23.6, I23.8, I24, I24.0-I24.1, I24.8-I24.9, I25.1-I25.2, I25.5-I25.6, I25.8-I25.9 |
| 41200/41210 (OPCS-4 surgical codes) | K40, K40.1-K40.4, I40.8-K40.9, K41, K41.1-K41.4, K41.8-K41.9, K42, K42.1-K42.4, K42.8-K42.9, K43, K43.1-K43.4, K43.8-K43.9, K44, K44.1-K44.2, K44.8-K44.9, K45.1-K45.6, K45.8-K45.9, K46, K46.1-K46.5, K46.8-K46.9, K49.1-K49.4, K49.8-K49.9, K50.1-K50.2, K50.4, K75.1-K75.4, K75.8-K75.9 |
| 41203/41205 (ICD-9 codes) | 410, 410.9, 411, 411.9, 4112, 412.9, 414.0, 414.8, 414.9 |

**Table S5 Cardiometabolic disease associations with BMI-unadjusted visceral, abdominal subcutaneous, and gluteofemoral adipose tissue volumes**

| | Type 2 Diabetes OR/SD (95% CI) | Coronary Artery Disease OR/SD (95% CI) |
|---|---|---|
| **Visceral adipose tissue** | 2.14 (2.05-2.23) | 1.35 (1.29-1.41) |
| **Abdominal subcutaneous adipose tissue** | 1.69 (1.63-1.75) | 1.23 (1.18-1.28) |
| **Gluteofemoral adipose tissue** | 1.48 (1.42-1.54) | 1.17 (1.12-1.22) |
| **Body mass index** | 1.87 (1.80-1.95) | 1.31 (1.26-1.37) |

Effect size estimates obtained from logistic regression models adjusted for age, sex, and MRI imaging center.

**Table S6 Standardized prevalence of type 2 diabetes across quintiles of VATadjBMI, ASATadjBMI, and GFATadjBMI.**

| T2D | VATadjBMI | | | ASATadjBMI | | | GFATadjBMI | | |
|---|---|---|---|---|---|---|---|---|---|
| | **Q1** | **Q2-Q4** | **Q5** | **Q1** | **Q2-Q4** | **Q5** | **Q1** | **Q2-Q4** | **Q5** |
| **Men** | | | | | | | | | |
| Normal | 1.6% (1.2 - 2.2%) | 2.8% (2.4 - 3.3%) | 6.6% (5.5 - 7.9%) | 1.9% (1.4 - 2.5%) | 2.8% (2.4 - 3.3%) | 3.5% (2.8 - 4.5%) | 5.2% (4.1 - 6.6%) | 2.8% (2.4 - 3.3%) | 1.8% (1.4 - 2.4%) |
| Over-weight | 2.7% (2.2 - 3.4%) | 4.9% (4.4 - 5.4%) | 9.6% (8.6 - 10.8%) | 3.8% (3.1 - 4.7%) | 4.9% (4.4 - 5.4%) | 6.1% (5.2 - 7.2%) | 8.4% (7.2 - 9.8%) | 4.9% (4.4 - 5.4%) | 3.3% (2.7 - 4.0%) |
| Obesity | 5.3% (4.3 - 6.4%) | 9.7% (8.6 - 10.9%) | 15.3% (13.3 - 17.7%) | 9.5% (7.9 - 11.4%) | 9.7% (8.6 - 10.9%) | 12.0% (10.2 - 14.2%) | 15.2% (13.0 - 17.7%) | 9.7% (8.6 - 10.9%) | 7.2% (5.8 - 8.7%) |
| **Women** | | | | | | | | | |
| Normal | 0.5% (0.4 - 0.8%) | 0.7% (0.5 - 0.8%) | 1.8% (1.4 - 2.4%) | 1.2% (0.9 - 1.7%) | 0.7% (0.5 - 0.8%) | 0.8% (0.6 - 1.2%) | 1.3% (1.0 - 1.8%) | 0.7% (0.5 - 0.8%) | 0.5% (0.4 - 0.8%) |
| Over-weight | 1.3% (0.9 - 1.7%) | 1.6% (1.3 - 1.9%) | 3.8% (3.2 - 4.6%) | 2.3% (1.8 - 2.9%) | 1.6% (1.3 - 1.9%) | 1.7% (1.3 - 2.2%) | 2.9% (2.4 - 3.6%) | 1.6% (1.3 - 1.9%) | 1.3% (1.0 - 1.7%) |
| Obesity | 3.5% (2.7 - 4.4%) | 4.5% (3.8 - 5.3%) | 9.2% (7.6 - 11.2%) | 5.1% (4.1 - 6.3%) | 4.5% (3.8 - 5.3%) | 4.0% (3.1 - 5.2%) | 7.6% (6.3 - 9.3%) | 4.5% (3.8 - 5.3%) | 3.6% (2.7 - 4.7%) |

Estimates were obtained from sex-specific logistic regressions adjusted for age, BMI, the other two fat depots, MRI imaging center and BMI by fat depot interaction terms. Covariates were set to sex- and BMI group-specific median age, median BMI, the Q2-Q4 fat depot strata (for the two fat depots not being varied) and mean MRI imaging center.

**Table S7 Standardized prevalence of coronary artery disease across quintiles of VATadjBMI, ASATadjBMI, and GFATadjBMI.**

| CAD | VATadjBMI | | | ASATadjBMI | | | GFATadjBMI | | |
|---|---|---|---|---|---|---|---|---|---|
| | Q1 | Q2-Q4 | Q5 | Q1 | Q2-Q4 | Q5 | Q1 | Q2-Q4 | Q5 |
| **Men** | | | | | | | | | |
| Normal | 5.2% (4.2 - 6.4%) | 5.5% (4.8 - 6.1%) | 6.7% (5.6 - 7.9%) | 5.0% (3.9 - 6.3%) | 5.5% (4.8 - 6.1%) | 6.1% (5.0 - 7.5%) | 6.6% (5.4 - 8.1%) | 5.5% (4.8 - 6.1%) | 4.4% (3.5 - 5.4%) |
| Overweight | 6.0% (5.2 - 7.0%) | 7.1% (6.5 - 7.7%) | 8.3% (7.4 - 9.3%) | 6.7% (5.7 - 7.8%) | 7.1% (6.5 - 7.7%) | 7.6% (6.6 - 8.7%) | 8.4% (7.3 - 9.7%) | 7.1% (6.5 - 7.7%) | 5.4% (4.7 - 6.3%) |
| Obesity | 6.9% (5.6 - 8.4%) | 9.2% (8.2 - 10.5%) | 10.4% (8.7 - 12.3%) | 9.1% (7.5 - 11.1%) | 9.2% (8.2 - 10.5%) | 9.3% (7.6 - 11.3%) | 10.9% (9.0 - 13.2%) | 9.2% (8.2 - 10.5%) | 6.8% (5.4 - 8.4%) |
| **Women** | | | | | | | | | |
| Normal | 1.0% (0.6 - 1.5%) | 1.1% (0.8 - 1.4%) | 1.4% (1.0 - 2.0%) | 1.4% (0.9 - 1.9%) | 1.1% (0.8 - 1.4%) | 1.1% (0.7 - 1.6%) | 1.1% (0.8 - 1.6%) | 1.1% (0.8 - 1.4%) | 1.0% (0.7 - 1.4%) |
| Overweight | 1.4% (1.0 - 1.9%) | 1.7% (1.4 - 2.0%) | 2.1% (1.6 - 2.6%) | 1.9% (1.5 - 2.5%) | 1.7% (1.4 - 2.0%) | 1.7% (1.3 - 2.2%) | 1.8% (1.4 - 2.3%) | 1.7% (1.4 - 2.0%) | 1.3% (0.9 - 1.7%) |
| Obesity | 1.9% (1.3 - 2.8%) | 2.4% (1.9 - 3.2%) | 2.8% (2.0 - 4.0%) | 2.5% (1.8 - 3.5%) | 2.4% (1.9 - 3.2%) | 2.5% (1.7 - 3.7%) | 2.8% (2.0 - 3.9%) | 2.4% (1.9 - 3.2%) | 1.5% (0.9 - 2.3%) |

Estimates were obtained from sex-specific logistic regressions adjusted for age, BMI, the other two fat depots, MRI imaging center, and BMI by fat depot interaction terms. Covariates were set to sex- and BMI group-specific median age, median BMI, the Q2-Q4 fat depot strata (for the two fat depots not being varied) and mean MRI imaging center.

**Table S8 Hazard ratios of VATadjBMI, ASATadjBMI, and GFATadjBMI for incident cardiometabolic disease**

| Type 2 Diabetes | | | |
|---|---|---|---|
| **No. Events / Total No. at Risk (%)** | **BMI-adjusted Fat Depot** | **HR (95% CI)** | **P value** |
| 235/37,891 (0.6) | VATadjBMI | 1.45 (1.30 - 1.61) | $1.3 \times 10^{-11}$ |
| | ASATadjBMI | 0.96 (0.84 - 1.08) | 0.49 |
| | GFATadjBMI | 0.84 (0.74 - 0.95) | 0.005 |
| **Coronary Artery Disease** | | | |
| **No. Events / Total No. at Risk (%)** | **BMI-adjusted Fat Depot** | **HR (95% CI)** | **P value** |
| 607/38,067 (1.6%) | VATadjBMI | 1.17 (1.08 - 1.26) | $8.1 \times 10^{-5}$ |
| | ASATadjBMI | 1.04 (0.95 - 1.14) | 0.41 |
| | GFATadjBMI | 0.91 (0.83 – 1.00) | 0.05 |

Hazard ratios with 95% CI in parentheses are shown for VATadjBMI, ASATadjBMI, and GFATadjBMI in Cox proportional-hazard models adjusted for age, sex, BMI, the other two fat depots, and MRI imaging center. Median follow-up time for both incident type 2 diabetes and coronary artery disease was 2.8 years from the date of imaging.
